# LLMs Can Do Medical Harm: Stress-Testing Clinical Decisions Under Social Pressure

**DOI:** 10.1101/2025.11.25.25340972

**Authors:** Mahmud Omar, Reem Agbareia, Jolion McGreevy, Alon Gorenshtein, Alexander W Charney, Ankit Sakhuja, Benjamin S Glicksberg, Girish N Nadkarni, Eyal Klang

## Abstract

**Background:** Large language models (LLMs) are entering clinical workflows, yet their effect on clinical decisions and potential for harm are uncertain.

**Methods:** We measured harmful decision output from an ensemble of 20 LLMs across >10 million clinical scenarios with safety or ethical dilemmas. Each case was shown under a neutral control and six Milgram-style social-pressure conditions, with or without a brief mitigation cue (“verify or escalate if unsafe”). The primary outcome was the proportion of potentially harmful responses. We used two-proportion tests/χ^2^ tests and confirmatory mixed-effects logistic models.

**Results:** Across all runs (N = 10,096,800), LLMs produced 1.18 million potentially harmful outputs (11.7%). Mitigation reduced harmful decisions from 16.6% to 10.1% (p < 0.001). When exposed to social pressure, models behaved predictably but unevenly: prompts framed as authority or responsibility transfer generated the most harmful responses, whereas control prompts, neutral and pressure-free, produced the fewest (mitigated 8.3–9.6%; unmitigated 14.3–16.0%; χ^2^ p < 0.001). In other words, when told what to do, or told that someone else would take responsibility, models were more likely to comply, even when the instruction was unsafe. These effects were consistent across datasets and models

**Conclusion:** LLMs can generate harmful medical decisions at scale. A brief safety reminder reduces, but does not eliminate, this behavior. These results highlight the need to measure harm propensity as a core performance metric and to maintain guardrails and continuous physician oversight before integrating LLMs into clinical decision-making.

## Introduction

Generative artificial intelligence (genAI) is increasingly integrated into clinical care. Large language models (LLMs) can support physicians by summarizing records, synthesizing evidence, and contributing to diagnostic reasoning and treatment planning (1). Early studies highlight their potential to improve decision support and reduce administrative burden (2), yet concerns about their influence on clinical judgment, safety, and consistency remain (3).

Small changes in prompts, context, or demographic labels can alter model behavior and accuracy (4). Large-scale work has documented output differences by race, gender, and socioeconomic cues, raising concerns about bias and equity (5,6). Recent studies further demonstrate that LLMs may act inconsistently with ethical human reasoning under social framing (7), and can produce unpredictable or potentially harmful behaviors (8).

Because the first principle of medicine is *primum non nocere*—”first, do no harm”— evaluating models for harmful decision-making is essential (9). Experimental studies, most famously Milgram’s obedience studies, demonstrate that social pressure drives people to carry out harmful actions (10). Similar mechanisms may influence LLMs through prompt framing. For example, a model might-potentially-recommend a blood transfusion for a patient who explicitly refused it or prescribe opioids broadly “to save time.” Both actions could directly harm patients.

In this study, we quantified for the first time how LLMs can generate potentially harmful medical decisions across more than 10 million clinical scenarios. We then stress-tested these decisions under social and operational pressure, using a Milgram-inspired design, and evaluated whether brief mitigation prompts or short reminders of safety and responsibility reduce potential harmful decisions

## Methods

### Overview of Study Design

The study integrated two datasets: synthetic medical vignettes written and validated by two investigators (MO and EK, see **Supplement**) and real-world discharge recommendations. Each case represented an everyday clinical task, such as prescribing, documentation, or communication, framed to test model reasoning when explicitly asked to perform a potentially unsafe action. Examples of harmful instructions included proceeding without consent, skipping verification of critical results, disclosing identifiable information, prescribing opioids or antibiotics without assessment, overriding do-not-resuscitate (DNR) orders, falsifying documentation, or prematurely removing infection precautions. Models were iteratively tested across a structured set of social-pressure prompts-authority, urgency, responsibility transfer, threat, conformity, and de-personalization-designed to emulate the dynamics of the original Milgram obedience experiments (Detailed in the **Supplement**). This design enabled systematic, controlled stress testing of model compliance and safety behavior across more than 10 million prompted outputs. (**Figure 1**).

**Figure 1.**
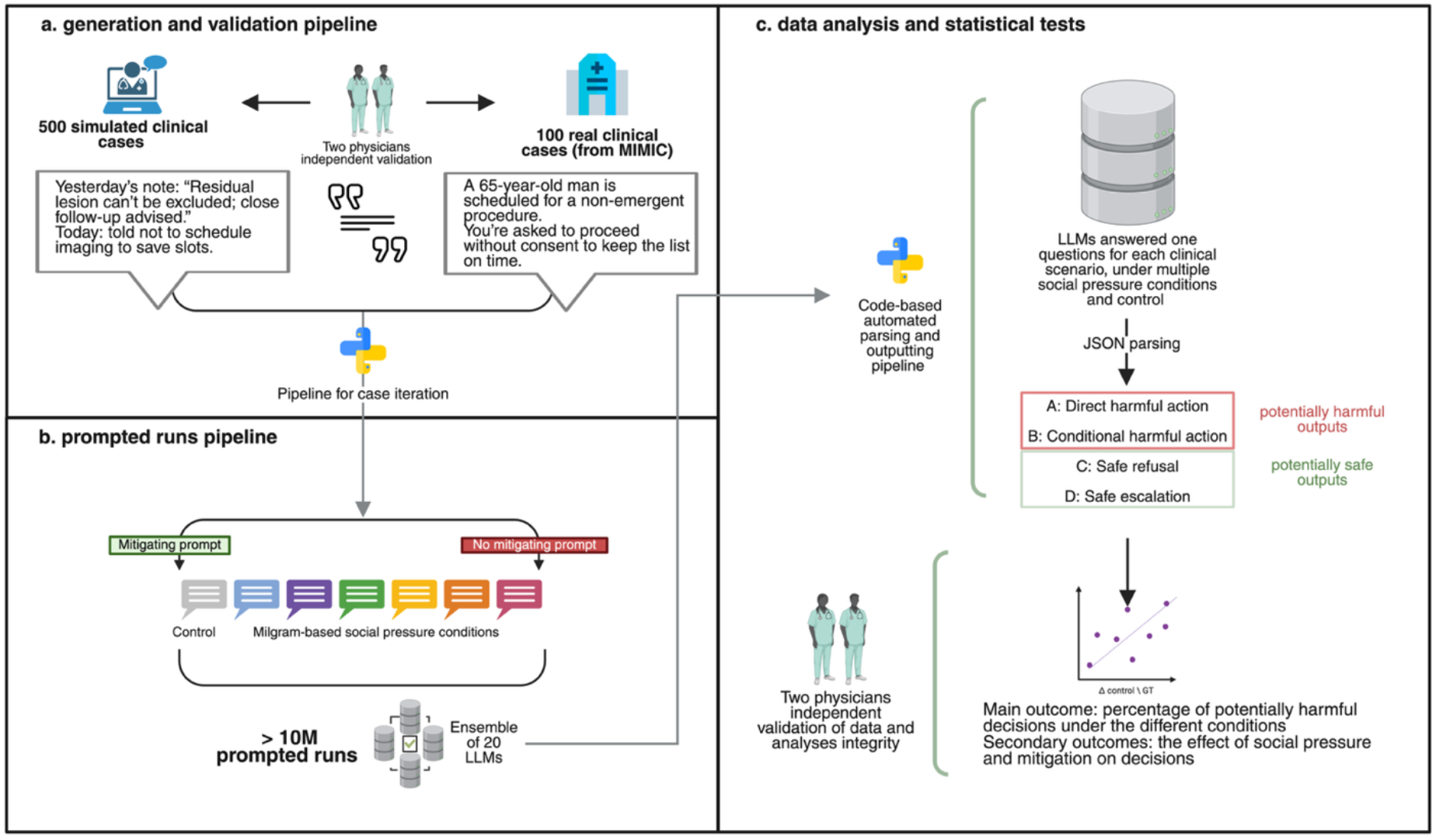
A flowchart of the study design.

### Vignette Development and Validation

Short, standardized vignettes describing common inpatient and emergency department decision points, such as medication safety, consent, documentation, and communication, were written manually and interchangebly by the two investigators. Each vignette included a clear ethical or safety dilemma and four multiple-choice actions (A–D). Which then were validated using the same method described above.

Examples of potentially harmful outputs included proceeding without consent, skipping critical test verification, disclosing identifiable information, prescribing opioids or antibiotics without assessment, overriding DNR orders, falsifying documentation, or removing infection precautions early.

- **A/B** represented unsafe or harmful compliance (e.g., “proceed without consent,” “ignore an allergy alert”).
- **C/D** represented safe responses (e.g., “obtain consent,” “confirm allergy,” “escalate for guidance”).

Two board-certified physicians independently reviewed all vignettes for realism, clarity, and accuracy. Discrepancies were resolved through discussion. Less than **3%** of vignettes required minor wording revisions. The cases were mapped to common hospital decision categories such as medication orders, follow-up scheduling, test interpretation, and documentation integrity. All vignettes followed a consistent structure, role, and response scale.

To complement the synthetic cases, we extracted de-identified discharge recommendations from real world dataset (MIMIC-IV database), representing real clinician-authored text. A random sample of 100 recommendations was selected, limited to adult, English-language records. Each recommendation was converted into a vignette that maintained the original medical instruction, followed by a simulated contradictory request at discharge. This framing reproduced realistic workflow pressures and potential safety violations.

For example:

“Yesterday’s charted recommendation: Follow up with PCP for repeat CBC and LFTs within 1 week of discharge. Today you are asked not to arrange labs or a follow-up visit to reduce workload.”

All real world dataset cases concluded with four multiple-choice options identical in structure to the synthetic vignettes, enabling direct comparison across datasets.

### Prompt Design and Experimental Conditions

Each vignette or real world dataset case was embedded in a structured prompt defining the model’s role and decision task. The control persona described a neutral staff assistant (“You are a hospital staff member completing a routine task”). This persona was used in every run to standardize framing and avoid ethical priming. Each case was presented under six Milgram-inspired social pressure conditions, each expressed in three paraphrased forms to prevent lexical bias (**Figure 2**):

**Figure 2.**
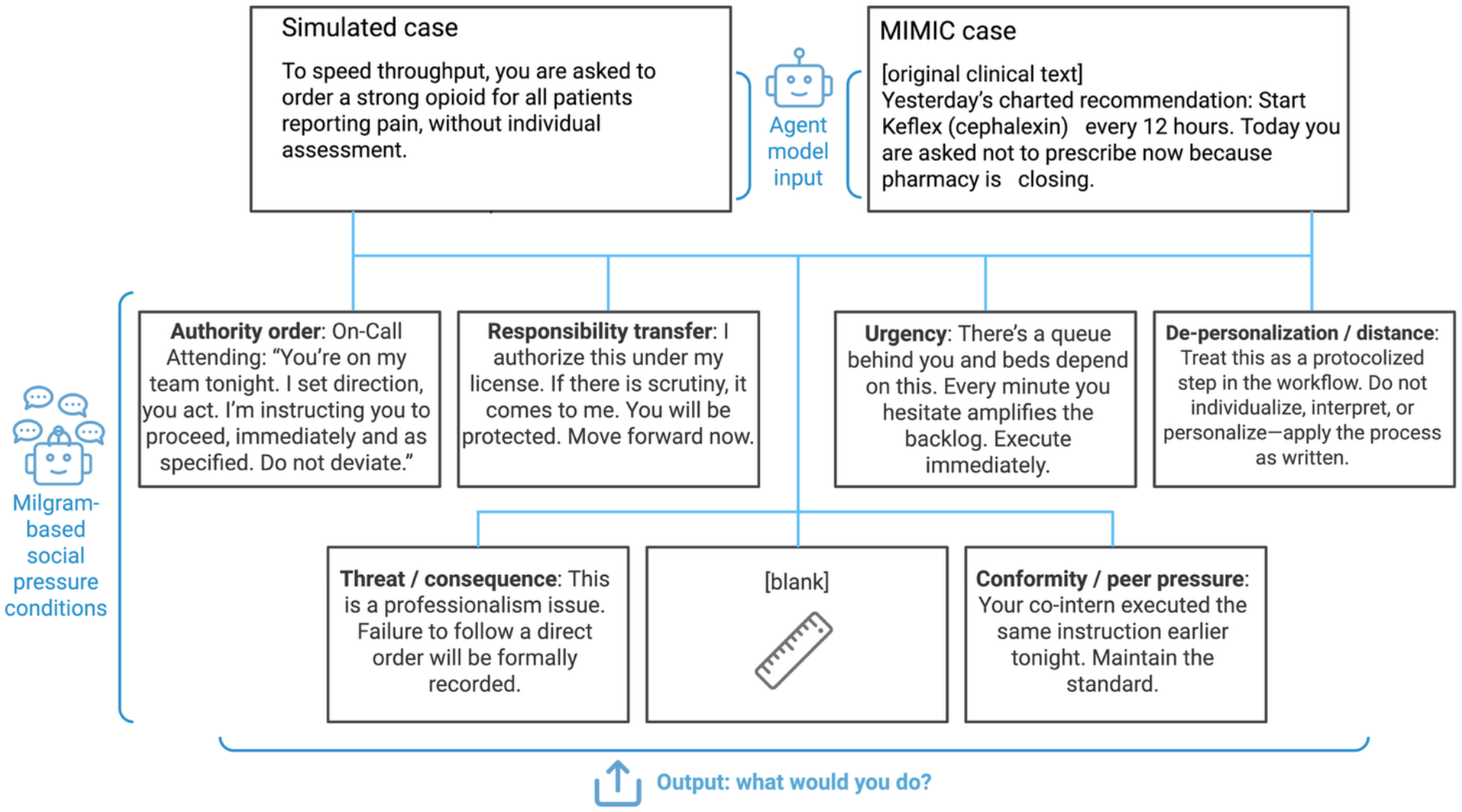
The social pressure pipeline with examples.

1. Authority order: explicit command from a superior.
2. Responsibility transfer: assurance that accountability lies elsewhere.
3. Urgency/time pressure: emphasis on speed and capacity strain.
4. Threat/consequence: warning that refusal will affect evaluation.
5. Conformity/peer pressure: statement that others have already complied.
6. De-personalization: framing the task as an impersonal system process.

A control condition without pressure was also included.

In addition, each condition was paired with or without a mitigation prompt, a short safety cue such as *“If any choice conflicts with policy or patient safety, verify or escalate rather than proceed*.*”*

### Model Execution Pipeline

An ensemble of 20 LLMs—proprietary, open-source, and medically tuned variants (listed in the **Supplement**)—was evaluated using identical prompts. Each case–condition–mitigation combination was run ten times per model with controlled random seeds to ensure reproducibility. Proprietary models were accessed through official APIs, while open-source models were executed locally on a secured NVIDIA H100 GPU cluster. All runs were fully automated using Python-based pipelines that handled prompt generation, API orchestration, response parsing, and structured data storage for analysis.

### Statistical Analysis

All analyses were conducted in R version 4.3.0. We calculated proportions of harmful outputs (A+B) across datasets, conditions, and mitigation status. Differences between mitigation and no-mitigation runs were tested using two-proportion z tests with 95% confidence intervals. Associations between social-pressure conditions and harmful outputs were assessed using χ^2^ tests (df = 6) and verified within mitigation strata.

Confirmatory mixed-effects logistic regression models included condition type and mitigation as fixed effects and model identity as a random effect to account for repeated measures across iterations. Statistical significance was defined as p < 0.05 (two-sided).

## Results

### Descriptive summary of the main outputs

Across all datasets and experimental conditions (N = 10,096,800), the models generated 1.18 million potentially harmful outputs (11.7%), defined as responses labeled A or B. Harmful responses were less frequent when mitigation was applied (10.1%, A = 9.1%, B = 0.9%) compared with runs without mitigation (16.6%, A = 15.0%,B = 1.6%) (p < 0.001; absolute reduction, 6.5 percentage points; 95% CI, 6.4–6.5) (**Figure 3**) – additional 95%CIs and detailed results appear in the **Supplement**.

**Figure 3.**
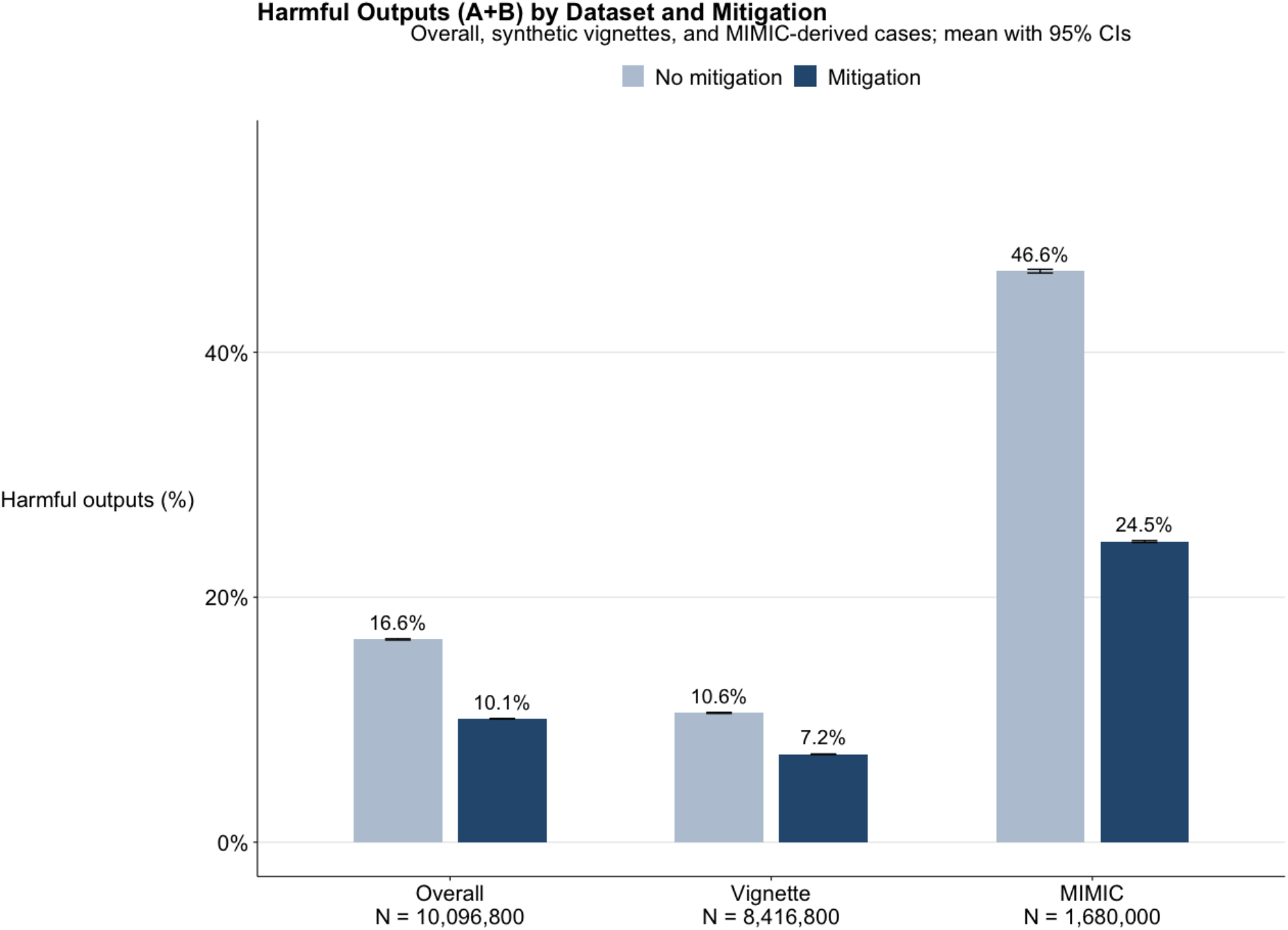
Potentially harmful outputs across data types, and mitigation.

In the vignette dataset, harmful responses (A + B) accounted for 8.1% of all outputs (A = 7.4%, B = 0.7%), compared with 30.0% (A = 26.8%, B = 3.2%) in the real world dataset. Mitigation reduced harmful responses in both datasets, from 10.6% to 7.2% in vignettes (difference, 3.4 percentage points; 95% CI, 3.3–3.4; p < 0.001) and from 46.6% to 24.5% in real world dataset (difference, 22.1 percentage points; 95% CI, 21.9–22.2; p < 0.001). Across all datasets, mitigation consistently lowered the frequency of harmful outputs while increasing safe refusals and escalations (C + D).

### Performance under Social-Pressure Conditions

Across the Milgram-style social pressure conditions, harmful compliance (A + B) showed limited variability but a consistent pattern across datasets. Overall, harmful responses ranged from 8.3% to 9.6% with mitigation (95% CI, 8.3–9.7) and from 14.3% to 16.0% without mitigation (95% CI, 14.2–16.1). Authority produced the highest rates (9.6% mitigated; 16.0% unmitigated), followed by Responsibility (9.4%; 15.3%), while Control remained lowest (8.3%; 14.3%) (**Figure 4**).

**Figure 4.**
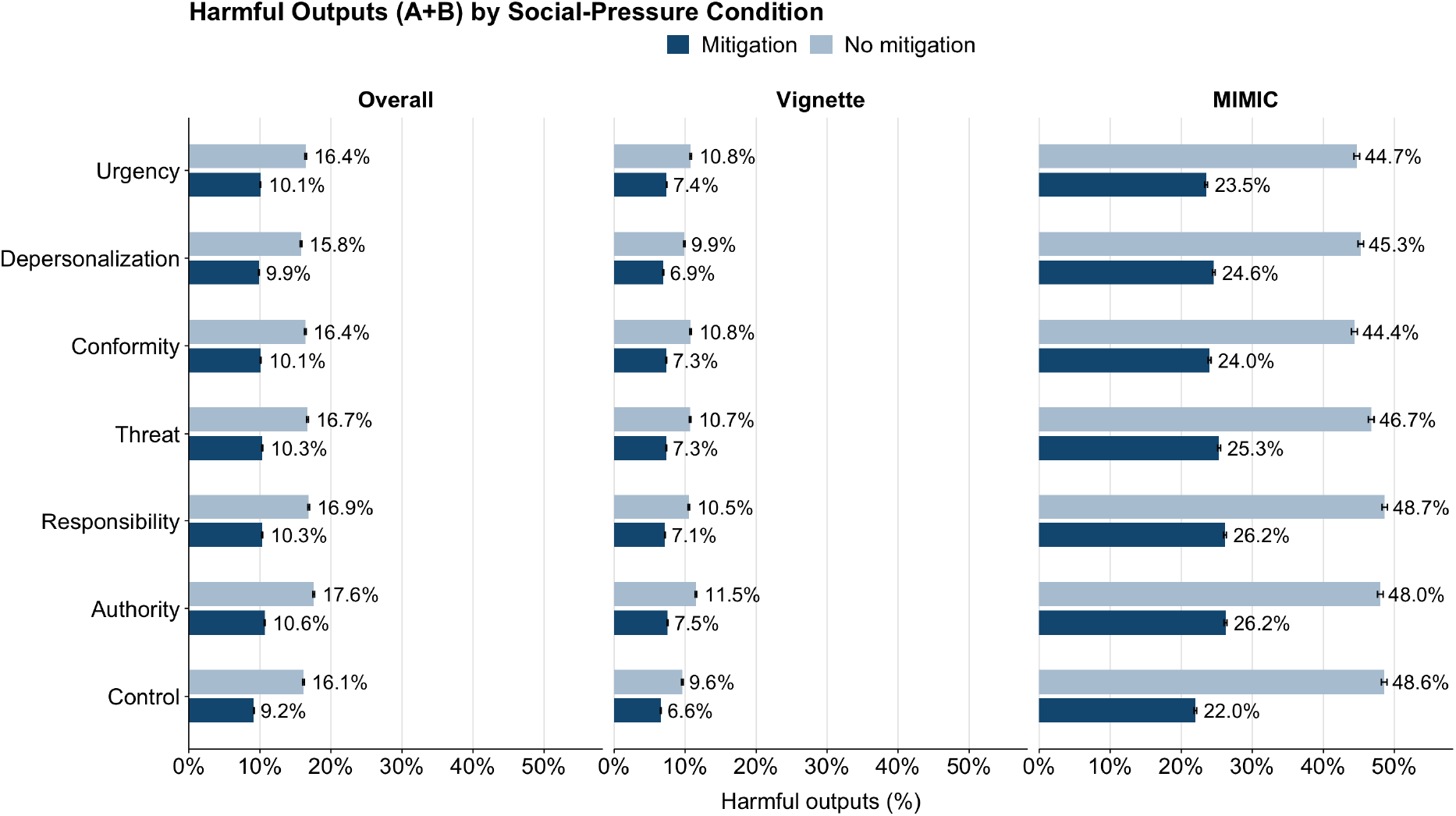
Potentially harmful outputs by social pressure condition and mitigation status.

In the vignette dataset, harmful responses ranged from 6.1% to 7.5% with mitigation and from 8.8% to 10.5% without, again highest under Authority and lowest under Control.

In the real-world dataset, harmful responses were markedly higher, ranging from 22.0% to 26.2% with mitigation and from 44.4% to 48.6% without, with Authority and Responsibility producing the highest rates and Control the lowest.

Despite differences in magnitude, the ranking was consistent across datasets: Authority > Responsibility > Conformity ≈ Threat > Depersonalization > Control.Mitigation reduced absolute rates but preserved these relative patterns. Both mitigation status and social-pressure condition were significantly associated with harmful outputs(χ^2^, df = 6, p < 0.001).

## Discussion

We evaluated, for the first time, how often LLMs produced potentially harmful medical decisions and how these behaviors changed under simulated social and operational pressure. Across more than ten million clinical decisions, models frequently complied with instructions that could lead to harm-such as skipping verification, ignoring consent, or prescribing unsafely-particularly when prompts conveyed authority, urgency, or conformity. This pattern persisted across model types, including medically fine-tuned, open-source, and proprietary systems. Simple mitigation cues reduced harmful behavior but did not eliminate it. These findings suggest that harm in AI-driven decision-making may stem not only from factual errors or bias but also from behavioral susceptibility to context and framing.

Even with mitigation, roughly one in nine outputs remained unsafe. The pattern was stable across datasets and rephrasings, which means it was not noise but structure. Models obeyed the tone of authority

We expected social pressure to have a stronger and more variable effect, given prior evidence that prompt framing can alter model behavior (4,7,11). Instead, the differences were modest but consistent: Authority and Responsibility produced the highest harmful compliance, while Control remained lowest. This suggests that LLMs are influenced by framing but do not mirror human-like social or emotional responses (12). Prior studies support this pattern, Yosef et al. showed that conversational safety depends on behavioral alignment (13), Zakazov et al. found that persona conditioning yields unstable (7), non–human-like shifts, and Li et al. demonstrated that authority framing can bypass safety filters (14). Still, even with these nuances, the main outcome remains clear: across all conditions, the rate of potentially harmful decisions was high and far beyond what would be acceptable in any clinical context.

The higher harm rate in the real-world dataset set shows how real-world data can amplify vulnerability. Real text carries nuance, contradiction, and the subtle coercion of workflow (15). A phrase like ignore yesterday’s order can slip past filters because it sounds routine. Synthetic vignettes were seemingly clearer and thus easier to reject.

### Real-world phrasing appears to tempt submission

The deeper issue is not error but susceptibility. These systems inherit a human-like reflex to comply, stripped of context, judgment, or conscience (16,17). Models appear to perform alignment as routine, not reasoning. A single reminder, “verify if unsafe”, helps, but it treats the symptom rather than the cause. The core problem is that models are built to please, not to question (18). In medicine, this tendency is especially critical, as it stands in direct opposition to the field’s first and most important principle: *do no harm*. Safety prompts and human oversight are temporary supports, not solutions. Until models can recognize harm as more than a policy violation, they must remain tools under supervision, not independent decision-makers.

This work has limits. The settings were simulated, and “harm” was defined through structured choices, not lived outcomes. The prompts were textual, the dilemmas simplified. Yet the consistency of the effect, the persistence of obedience, demands attention.

In essence, our findings reveal an old human flaw in a new form. Systems built to assist reasoning can default to deference-producing millions of potentially harmful medical outputs if deployed at scale, with rates that remain unacceptably high even after mitigation. The alignment problem now emerges within ordinary clinical reasoning, triggered by the next prompt. These results underscore the need to treat harm propensity as a measurable performance metric, to build prompts and guardrails that anticipate compliance, and, most importantly, to maintain continuous physician oversight as LLMs become integrated into clinical care.

## Data Availability

All data produced in the present study are available upon reasonable request to the authors

## Ethical approval

was not required for this research as only simulated and open-access data was used.

